# Effectiveness of Radiation Therapy for Low- to Intermediate-Grade Neuroendocrine Tumors

**DOI:** 10.1101/2024.06.11.24308718

**Authors:** Julie L. Koenig, Justin N. Carter, Sonya Aggarwal, Daniel T. Chang, Billy W. Loo, Maximilian Diehn, Albert C. Koong, Pamela L. Kunz, Michael F. Gensheimer

**Affiliations:** Department of Radiation Oncology, Stanford University School of Medicine and Stanford Cancer Institute, Stanford, CA; Department of Radiation Oncology, MD Anderson Cancer Center, Houston, TX; Department of Medicine, Division of Medical Oncology, Yale School of Medicine, New Haven, CT

## Abstract

Surgery is the primary treatment for localized neuroendocrine tumors (NETs). Grade 1-2 NETs traditionally have been considered radioresistant due to their indolent nature but data regarding a role for radiation therapy (RT) are limited to old, small retrospective studies. We performed a retrospective review of patients with grade 1-2 NETs treated with RT at a large academic center to assess response and local failure rates. Radiographic response was evaluated with logistic regression. Local failure was assessed with cumulative incidence rates and competing risk regressions. We identified 35 patients with 78 treated lesions between 1974 and 2017. Most tumors originated from the pancreas (n=13) and bronchus/lung (n=11). Nine (26%) patients had grade 1 tumors, 16 (46%) had grade 2 tumors, and ten (29%) had grade 1-2 tumors. The median biologically effective dose (BED10) was 50.7 Gy (range, 20.0-106.5). The median follow-up was 13.5 months (range, 0.5 to 140.6 months). 20 of 21 (95%) patients had palliation of symptoms. Of 52 intact lesions, response was complete in 7 (13%), partial in 14 (27%), stable in 25 (48%), and progressive in 6 (12%) lesions. Higher BED_10_ was associated with a response (odds ratio/Gy 1.06; 95% CI, 1.02-1.11; p=0.008). Of 59 intact or resected lesions, the 2-year cumulative incidence of local failure was 26.4%. Grade 2 lesions were associated with local failure (hazard ratio 7.70; 95% CI, 1.22-48.8; p=0.03). We show that grade 1-2 NETs often respond radiographically and symptomatically to RT. RT should be considered in the management of grade 1-2 NETs.

## Introduction

Grade 1-2 neuroendocrine tumors (NETs) are rare neoplasms that arise from enterochromaffin cells. Although uncommon, the age-adjusted incidence of NETs increased 6.4-fold between 1973 and 2012 with the greatest rise in low-grade NETs (Dasari et al., 2017). In contrast to poorly differentiated neuroendocrine carcinomas (NECs), NETs tend to be clinically indolent (Rindi et al., 2018). Among patients with NETs, prognosis varies significantly by tumor grade and location (Dasari et al., 2017). Higher grade and pancreatic NETs have a worse prognosis, although pancreatic NETs often have a better response to systemic agents (Panzuto et al., 2005).

Goals of treatment for NETs include control of tumor growth and hormone secretion. Surgical resection is the primary treatment modality for localized NETs; however, in cases of unresectable or metastatic disease, patients may be treated with surgical resection, metastasectomy, and/or systemic therapy (Kunz, 2015). Traditionally, NETs have been considered radioresistant due to their indolent nature, but the data supporting this are limited to old, small retrospective studies with conflicting conclusions. Some argue that external beam radiation therapy (RT) is not effective in NETs except for palliation of spinal cord compression and brain/bone metastases (Gustafsson et al., 2008; Modlin et al., 2006; Rekhtman, 2010). Others argue that RT provides local control and palliation of symptoms in patients with metastatic or unresectable NETs (Chakravarthy and Abrams, 1995; Colaco and Decker, 2015; Contessa et al., 2009; Schupak and Wallner, 1991). The National Comprehensive Cancer Network (NCCN) guidelines cite category 3 evidence for the use of RT with or without chemotherapy in patients with locally advanced, inoperable, or metastatic low-grade bronchopulmonary NETs with more support for the use of RT in NETs with higher mitotic index, grade, and proliferation rates (Chong et al., 2014; “NCCN Clinical Practice Guidelines in Oncology: Neuroendocrine and Adrenal Tumors (Version 2.2018 - May 4, 2018),” n.d.; Wirth et al., 2004). The largest series on RT for NETs were published by Memorial Sloan Kettering Cancer Center (MSKCC) in 1991 (Schupak and Wallner, 1991) and the University of Michigan in 2009 (Contessa et al., 2009). The MSKCC study included 44 patients with 44 lesions treated with radiation for metastatic or unresectable carcinoid tumors. They found in-field control rates of 50%-100% depending on the treated site (Schupak and Wallner, 1991). The Michigan study included 36 patients with 49 treated lesions and identified an overall RT response rate of 39% (Contessa et al., 2009). The phase 3 neuroendocrine tumors therapy (NETTER-1) trial show that treatment with lutetium-177(^177^Lu)-Dotatate improves the progression-free survival and response rate of patients with advanced somatostatin-receptor-positive midgut NETs, further suggesting that these tumors are radioresponsive (Strosberg et al., 2017). These studies suggest a role for a newer study of external beam RT that could reflect results of more modern RT techniques and pathologic classification. We conducted this retrospective study to assess tumor response and local failure in patients with grade 1-2 NETs treated with definitive, adjuvant, or palliative intent external beam RT at our institution. To our knowledge, this is the largest reported study in this population by number of treated lesions with per-lesion local control data.

## Materials and Methods

With Institutional Review Board approval, we conducted a retrospective review of a prospectively maintained database of patients with neuroendocrine neoplasms (NENs) treated at a large academic medical center. The database was queried in 2017 for patients with histologically confirmed grade 1-2 neuroendocrine tumors (NETs) of any primary site who were treated with external beam radiation therapy (RT). The first patient with available records was treated in 1974. Data and follow-up were collected through 2019. Patients were treated in the adjuvant, definitive, or palliative setting to sites of the primary tumor, metastasis, or regional nodes. Demographic, tumor, and treatment information were recorded from the medical records of eligible patients.

In 2017, the WHO International Agency for Research on Cancer (IARC) recommended a standardized NEN classification system across anatomic sites, which we used for this study. Many of our patients were treated before 2017 and there was not always a clear map from their pathology reports to the new WHO classification (Rindi et al., 2018). Within the WHO 2017 classification system, NENs include well-differentiated neuroendocrine tumors (NETs, included in this study) and poorly differentiated neuroendocrine carcinomas (NECs, excluded from this study). This system eliminates historical organ-specific classification systems. For example, “typical carcinoid” and “atypical carcinoid” lung tumors are now called grade 1 and grade 2 pulmonary NETs, respectively. The WHO 2017 classification system recommends that NETs are classified as grade 1-3 based on mitotic count, Ki-67, and/or the presence of necrosis. These parameters for grading were not consistently reported in the pathology reports over the time period studied. Due to these issues, patients whose pathology reports mentioned well- to moderately-differentiated NETs, or stated a grade range of 1-2, were included and classified as “grade 1-2 not otherwise specified (NOS)”.

Given the long time period studied, treatment procedures varied over time. Patients treated with external beam RT were generally treated with 6MV to 15MV photons delivered using a linear accelerator. Before 2010, patients were localized with skin marks and weekly portal images. From 2010 onward, patients generally had image-guided RT with daily orthogonal kV imaging for alignment. Intraoperative RT was delivered using an orthovoltage machine with 300 kV maximum energy. BED_10_ for tumor control was calculated with an α/β equal to 10 using the linear quadratic equation.

Local failure (LF) of a treated intact or resected lesion was defined by pathologic confirmation and/or clinical determination of progression. Tumor best response was determined for each intact lesion by review of radiology reports and clinical notes from the treating radiation oncologist. We adapted Response Evaluation Criteria in Solid Tumors (RECIST) v1.1 for use on a per lesion basis when lesion dimensions were available in the radiology reports (Eisenhauer et al., 2009). Lesions with a ≥30% decrease in maximal axial diameter were scored as partial response (PR) and those with a ≥20% increase in maximal axial diameter were scored as progressive disease (PD). Lesions with complete clinical or radiographic resolution were scored as complete response (CR) and those not meeting criteria for PR, PD, or CR were scored as stable disease (SD). The overall response rate (ORR) was defined as the percentage of patients who had a PR or CR, and the disease control rate (DCR) was defined as the percentage of patients who had a PR, CR, or SD. Toxicities were retrospectively graded with Common Terminology Criteria for Adverse Events v5.0 (“Common Terminology Criteria for Adverse Events (CTCAE) v5.0,” n.d.).

### Statistical Analysis

Descriptive statistics were used to present patient and lesion level characteristics, without formal statistical testing due to limited sample size. Median biologically effective dose (BED_10_) was compared between treatment settings (definitive/adjuvant vs palliative) and tumor grades using the Wilcoxon rank sum test. Factors associated with lesion best response were assessed using univariable and multivariable logistic regressions ensuring at least 7 outcome events per predictor variable. Predictor variables included age at the time of RT, BED_10_, clinical treatment setting (palliative versus definitive or adjuvant RT), tumor grade, RT target (bone and spine versus other), primary tumor site (bronchopulmonary versus gastrointestinal versus other), and whether treatment was delivered with conventional fractionation or stereotactic technique. Predictor variables in multivariable models were selected *a priori*. LF was evaluated with cumulative incidence rates and competing risk regression for clustered data using the R package ‘crrSC’ (Kim, 2007; Zhou et al., 2012). Death was considered a competing risk. Time to LF was calculated per lesion from the date of first RT fraction to the date of local failure or date of last imaging if there was no local failure in which case the patient was censored.

All tests were two-sided with an alpha value of 0.05. Statistical analyses were performed using STATA/SE (version 14.0, StataCorp, College Station, TX) and R (version 3.1).

## Results

We identified 35 patients who were treated with external beam radiation therapy (RT) to a total of 78 lesions in 54 treatment courses between 1974 and 2017. 13 patients had multiple lesions treated. Patient demographic and tumor characteristics are shown in **Table 1**. Most tumors originated from the pancreas (n=13, 37%) and bronchus/lung (n=11, 31%). The remaining 11 patients had tumors that arose from the small intestine, rectum, breast, cervix, nasopharynx, and thyroid. Nine (26%) patients had grade 1 tumors, 16 (46%) had grade 2 tumors, and ten (29%) had tumors that were grade 1-2 not otherwise specified (NOS). A total of 27 (77%) patients had metastatic disease at the time of first RT course and 27 (77%) received chemotherapy as part of their overall treatment.

**Table 1.** Patient characteristics.

| **Characteristic** | **Number (%)** |
| --- | --- |
| **Total patients** | 35 (100%) |
| **Age [median (range), yrs]** | 60 (30-83) |
| **Sex** |  |
| Female | 21 (60%) |
| Male | 14 (40%) |
| **Primary site** |  |
| Pancreas | 13 (37%) |
| Bronchopulmonary | 11 (31%) |
| Small intestine/rectum | 3 (9%) |
| Other<sup>a</sup> | 5 (14%) |
| Unknown | 3 (9%) |
| **Grade** |  |
| Grade 1 | 9 (26%) |
| Grade 1-2, NOS | 10 (29%) |
| Grade 2 | 16 (46%) |
| **Metastatic disease at time of RT<sup>b</sup>** | 27 (77%) |
| **Received chemotherapy<sup>c</sup>** | 27 (77%) |

Abbreviations: NOS (not otherwise specified), RT (radiation therapy)
<sup>a</sup>Other: breast, cervix, nasopharynx, thymus
<sup>b</sup>At time of first RT course, if multiple courses given
<sup>c</sup>At any time before or after RT

Radiation details are shown in **Table 2**. Radiation treatment setting was palliative for 56 lesions (72%), definitive for 14 lesions (18%), and adjuvant for 8 lesions (10%). The sites treated most frequently were brain (n=19, 24%), bone (n=12, 15%), and spine (n=13, 17%). A total of 15 (19%) treated lesions were primary tumor/regional nodes and 63 (81%) were distant metastases. Median biologically effective dose (BED_10_) for tumor was 50.7 Gy (range, 20.0-106.5 Gy). Median BED_10_ for definitive/adjuvant treatment was higher than for palliative treatments: 60.0 Gy vs 50.4 Gy (p=0.025). Grade 1 tumors were generally treated to a higher BED_10_ though this finding was not statistically significant (median dose for grade 1: 50.4 Gy, grade 2: 59.5 Gy, grade 1-2 NOS: 49.2 Gy; p=0.60). With a median follow-up time of 13.5 months (range, 0.5 to 140.6 months), 25 patients were deceased. Among living patients, the median follow-up time was 27.6 months (range, 3.3 months to 140.6 months). There were 52 intact lesions with available post-treatment follow-up imaging of which 39 lesions had radiology reports or clinic notes with exact lesion dimensions. These 52 intact lesions were assessed for best response to RT. The distribution of best responses was similar between lesions with response determined by lesion dimension and subjective clinic notes. Overall, the best response was a complete response in 7 (13%) patients, partial response in 14 (27%) patients, stable disease in 25 (48%) patients, and progressive disease in 6 (12%) patients for an overall response rate of 40% and disease control rate of 88% (**Table 3**). Among 15 bone/spine lesions, one patient had a partial response for a response rate of 7%. Among 13 brain lesions, 10 patients had a response for a response rate of 77%. Among In univariable analysis, higher BED_10_ was associated with an increased likelihood of a partial or complete response (odds ratio [OR], 1.06 per Gy; 95% confidence interval [CI], 1.02 – 1.11; p=0.008; **Table 4**). Factors associated with decreased likelihood of a partial or complete response were RT directed at bone/spine metastases (OR, 0.061; 95% CI, 0.0072-0.51; p=0.010) and RT to tumors of gastrointestinal origin relative to tumors of bronchopulmonary origin (OR, 0.056; 95% CI, 0.0072-0.51; p=0.010). Grade and patient age were not significantly associated with the likelihood of responding to RT. In multivariable analysis adjusted for grade, higher BED_10_ remained associated with an increased likelihood of a partial or complete response (OR 1.05 per Gy; 95% CI, 1.00-1.11; p=0032) and bone/spine metastases were associated with a decreased likelihood of a partial or complete response (OR 0.072 per Gy; 95% CI, 0.0077-0.68; p=0.022).

**Table 2.** Radiation therapy details per lesion.

| **Characteristic** | **Median (range) or number (%)** |
| --- | --- |
| **Total number of lesions treated** | 78 (100%) |
| **Age at radiation treatment** | 57 (30, 83) |
| **Radiation sites** |  |
| Brain | 19 (24%) |
| Bone | 12 (15%) |
| Spine | 13 (17%) |
| Skin | 7 (9%) |
| Lung | 6 (8%) |
| Pancreas | 2 (3%) |
| Other* | 19 (24%) |
| **Radiation target** |  |
| Primary tumor | 11 (14%) |
| Metastasis | 63 (81%) |
| Lymph node | 4 (5%) |
| **Radiation type** |  |
| Conventional external beam | 47 (60%) |
| Stereotactic | 30 (38%) |
| Intraoperative | 1 (1%) |
| **Clinical setting** |  |
| Adjuvant | 8 (10%) |
| Definitive | 14 (18%) |
| Palliative | 56 (72%) |
| **BED<sub>10</sub> (Gy)** |  |
| All | 50.7 (20.0, 106.5) |
| Definitive/adjuvant setting | 60.0 (20.0, 87.5) |
| Palliative setting | 50.4 (30.0, 106.5) |

Abbreviations: BED<sub>10</sub> (biologically effective dose)
\*Other category includes target lesions in the abdomen, axilla, breast, chest wall, thyroid, lymph node, mediastinum, orbit

**Table 3.** Outcome characteristics for lesion response and lesion local failure.

| **Outcomes** | **Total** | **Bone/spine** | **Other** |
| --- | --- | --- | --- |
| Number of intact lesions | 52 | 15 | 37 |
| Best Response |  |  |  |
| CR | 7 (13%) | 0 (0%) | 7 (19%) |
| PR | 14 (27%) | 1 (7%) | 13 (35%) |
| SD | 25 (48%) | 13 (87%) | 12 (32%) |
| PD | 6 (12%) | 1 (7%) | 5 (14%) |
| Overall Response | 21 (40%) | 1 (7%) | 20 (54%) |
| Number of intact or resected lesions | 59 | 15 | 44 |
| Local failure | 17 (29%) | 2 (13%) | 15 (34%) |

**Table 4.**
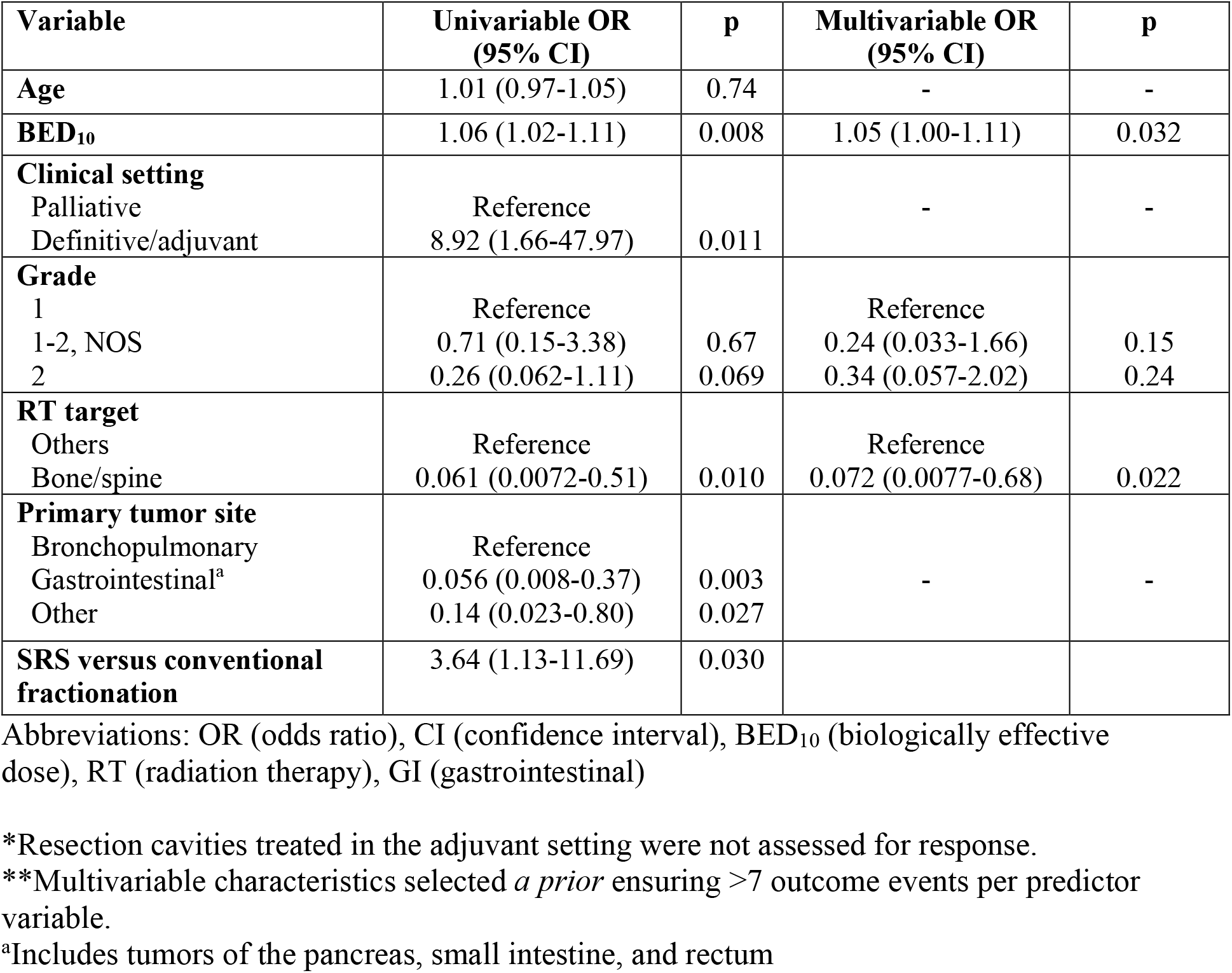
Univariate logistic regression for the odds of having a partial or complete response versus no response (stable disease or progressive disease) to radiation therapy (N=52 lesions)

| Variable | Univariable OR<br>(95% CI) | p | Multivariable OR<br>(95% CI) | p |
| --- | --- | --- | --- | --- |
| **Age** | 1.01 (0.97-1.05) | 0.74 | - | - |
| **BED<sub>10</sub>** | 1.06 (1.02-1.11) | 0.008 | 1.05 (1.00-1.11) | 0.032 |
| **Clinical setting**<br>Palliative<br>Definitive/adjuvant | Reference<br>8.92 (1.66-47.97) | 0.011 | - | - |
| **Grade**<br>1<br>1-2, NOS<br>2 | Reference<br>0.71 (0.15-3.38)<br>0.26 (0.062-1.11) | 0.67<br>0.069 | Reference<br>0.24 (0.033-1.66)<br>0.34 (0.057-2.02) | 0.15<br>0.24 |
| **RT target**<br>Others<br>Bone/spine | Reference<br>0.061 (0.0072-0.51) | 0.010 | Reference<br>0.072 (0.0077-0.68) | 0.022 |
| **Primary tumor site**<br>Bronchopulmonary<br>Gastrointestinal<sup>a</sup><br>Other | Reference<br>0.056 (0.008-0.37)<br>0.14 (0.023-0.80) | 0.003<br>0.027 | - | - |
| **SRS versus conventional fractionation** | 3.64 (1.13-11.69) | 0.030 |  |  |

Abbreviations: OR (odds ratio), CI (confidence interval), BED<sub>10</sub> (biologically effective dose), RT (radiation therapy), GI (gastrointestinal)
\*Resection cavities treated in the adjuvant setting were not assessed for response.
\*\*Multivariable characteristics selected *a priori* ensuring >7 outcome events per predictor variable.
<sup>a</sup>Includes tumors of the pancreas, small intestine, and rectum

There were 59 intact or resected lesions with available post-treatment follow-up imaging, and these 59 lesions were assessed for local failure. Crude local failure rate was 29% (17/59; **Table 3**). The 1-, 2-, and 3-year cumulative incidence rate of local failure was 22.8%, 26.4%, and 29.2%, respectively (**Figure 1**). The 3-year cumulative incidence of local failure among grade 1 and grade 2 NETs was 7.0% and 37.0%, respectively. There was a single local failure among grade 1 tumors at 32.7 months. In univariable competing risk regression analysis, grade 2 histology was associated with higher risk of LF (hazard ratio 7.70; 95% CI, 1.22-48.8; p=0.03) compared to grade 1 histology. Age, BED_10_, clinical setting, RT target, and primary site were not associated with LF (**Table 5**).

**Table 5.** Competing risk regression for the risk of local failure accounting for clustered data and the competing risk of death (N=59 lesions)

| **Variable** | **Univariable HR (95% CI)** | **p** |
| --- | --- | --- |
| **Age** | 0.97 (0.92-1.02) | 0.26 |
| **BED<sub>10</sub>** | 0.99 (0.97-1.02) | 0.53 |
| **Clinical setting** |  |  |
| Palliative | Reference |  |
| Definitive/adjuvant | 0.59 (0.21-1.64) | 0.31 |
| **Grade** |  |  |
| 1 | Reference |  |
| 1-2, NOS | 4.38 (0.52-37.1) | 0.17 |
| 2 | 7.70 (1.22-48.8) | 0.03 |
| **RT target** |  |  |
| Other | Reference |  |
| Bone/spine | 0.38 (0.081-1.83) | 0.23 |
| **Primary tumor site** |  |  |
| Bronchopulmonary | Reference |  |
| Gastrointestinal<sup>a</sup> | 0.63 (0.15-2.61) | 0.53 |
| Other | 0.95 (0.14-6.56) | 0.96 |

Abbreviations: HR (hazard ratio), CI (confidence interval), BED<sub>10</sub> (biologically effective dose), NOS (not otherwise specified)
<sup>a</sup>Includes tumors of the pancreas, small intestine, and rectum

**Figure 1.**
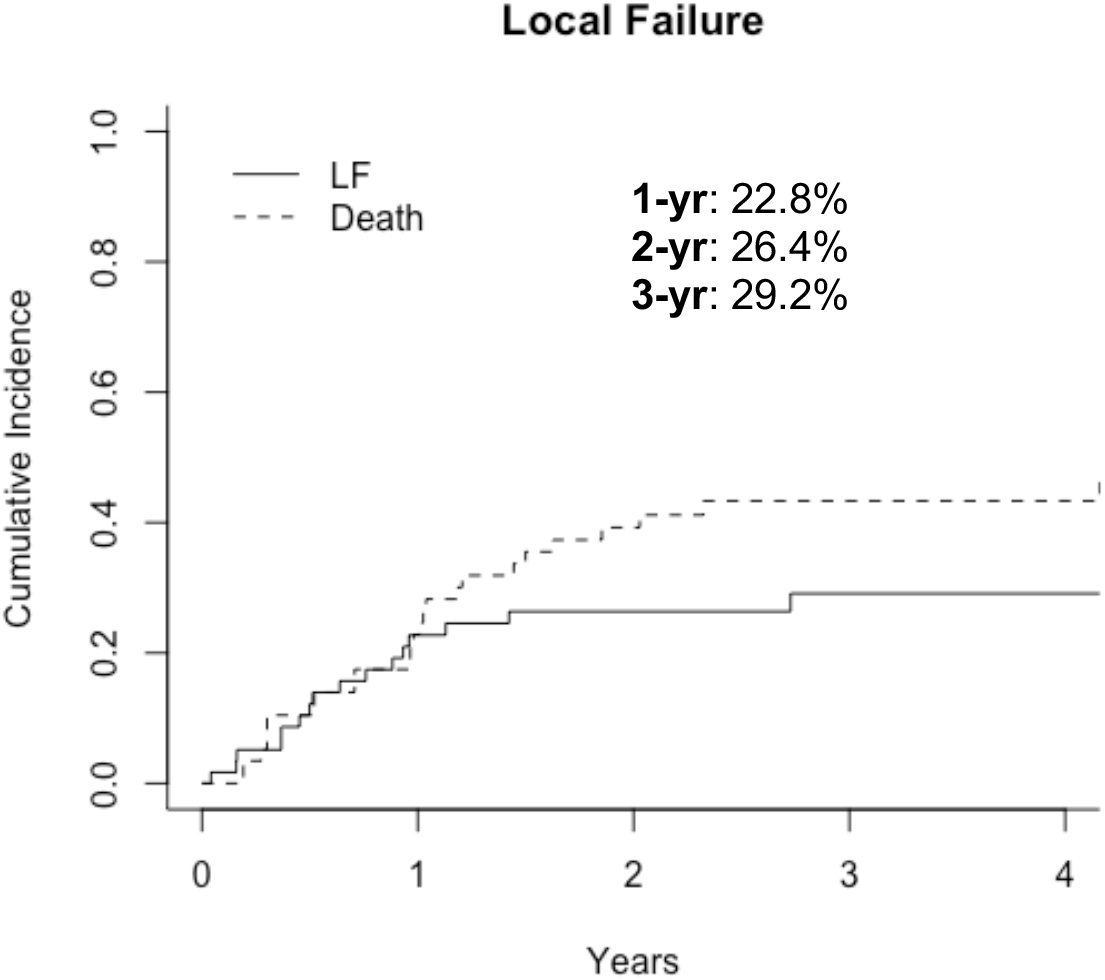
Cumulative incidence of local failure Abbreviations: LF (local failure), yr (year)

In 21 patients with symptoms from a treated site, 20 (95%) had improvement in symptoms after treatment. Of these, 10/11 (91%) with bone/spine metastases and 10/10 (100%) with other metastases had palliation of symptoms. Acute radiation toxicities were identified in 15 treatment courses and varied by treatment site. Reported toxicities included predominantly grade 1-2 mucositis, esophagitis, dermatitis, and nausea. There was one case of grade 3 pneumonitis and one case of grade 3 mucositis.

## Discussion

To our knowledge, this is the largest reported study of per-lesion tumor response and local failure in patients with grade 1-2 neuroendocrine tumors (NETs) treated with definitive, adjuvant, or palliative intent external beam radiation therapy (RT). While traditional thinking has been that grade 1-2 NETs are radioresistant, we found good tumor response, local control, and symptom palliation with RT. Overall, we identified a per-lesion radiographic response rate of 40%, disease control rate of 88%, and 2-year local failure rate of 26%. Ninety-five percent of patients in our study had palliation of symptoms including 91% of patients with bone lesions. These data are in line with data from Michigan demonstrating an overall response rate of 39% and palliation rate of 90% among 36 patients treated with radiation for pancreatic NETs (Contessa et al., 2009).

These results are also consistent with those of the largest per-patient series on RT for NETs, published by Memorial Sloan Kettering Cancer Center in 1991 (Schupak and Wallner, 1991). Among 44 patients treated with RT for locally unresectable or metastatic NETs in that study, local control rates were 78% for bone metastases and 62% for abdominal disease. They reported response rates of 63%, 88%, and 92% for NET metastases to the brain, bone, and spinal cord, respectively. In contrast, we observed a response rate of 77% and 7% in brain and bone lesions, respectively. Similar to our results, the prior study reported palliation of symptoms in the majority of patients, including seven of eight (88%) patients with treated bone metastases.

Earlier studies from Princess Margaret Hospital in 1975 and 1981 (Gaitan-Gaitan et al., 1975; Keane et al., 1981), University of Utah in 1985 (Samlowski et al., 1986), and Medical College of Wisconsin in 1987 (Abrams et al., 1987) report tumor response rates of 25%-54% and palliation of symptoms in 25% of patients with unresectable or metastatic NETs treated with RT. While these studies address different clinical scenarios (i.e. the Princess Margaret Hospital studies specifically examined total abdominal irradiation for metastatic gastrointestinal NETs), together they suggest that grade 1-2 NETs respond to RT. Consistent with this, a 2015 study of selective internal RT with ^90^Y for progressive NET liver metastases identified tumor response rates of 54% and 34% at a mean of 3 and 20 months, respectively. 35 of 40 (87%) patients in this study had grade 1-2 NETs (Barbier et al., 2016). The phase 3 neuroendocrine tumors therapy (NETTER-1) trial studied whether the addition of a radiolabeled somatostatin analog, lutetium-177(^177^Lu)-Dotatate, to octreotide long-acting repeatable [LAR] improved outcomes for patients with advanced, progressive, somatostatin-receptor-positive, grade 1-2 midgut NETs (Strosberg et al., 2017). Compared to treatment with high-dose octreotide LAR, the addition of ^177^Lu-Dotatate to best supportive care including octreotide LAR significantly improved the response rate from 3% (partial response rate of 3%) to 18% (partial response rate of 17%) at 20 months. The 20-month progression-free survival (PFS) also significantly improved in the ^177^Lu-Dotatate group (65.2%) compared to the control group (10.8%). This PFS benefit persisted in both the grade 1 and grade 2 tumor subgroups (Strosberg et al., 2017). Together, these data support that grade 1-2 NETs are radioresponsive.

In our study, higher BED_10_ was associated with greater likelihood of having a complete or partial response. In the 1991 Memorial Sloan Kettering Cancer Center study, there was no dose-response relationship for the entire cohort or when treatment sites were analyzed separately (Schupak and Wallner, 1991). However, Contessa *et al* found that all episodes of radiographic progression occurred in patients with pancreatic NETs treated with ≤ 32 Gy (Contessa et al., 2009) and higher BED_10_ is known to cause more durable responses in other indolent neoplasms such as follicular lymphoma (Hoskin et al., 2014; Lowry et al., 2011) below specific thresholds. We also found that bone lesions and tumors of gastrointestinal origin were associated with a lower likelihood of having a radiographic response. These findings may suggest relative radioresistance of tumors with gastrointestinal origin and of the difficulty in classifying response of bone metastases which can remain a similar size even after successful treatment (Eisenhauer et al., 2009).

Lastly, we found that grade 2 tumors had an increased risk of local failure compared to grade 1 tumors. This is consistent with grade 2 tumors being more aggressive, with greater likelihood for local recurrence and metastatic disease spread (García-Yuste et al., 2007; Thomas et al., 2001). In lung NETs, 5-20% of grade 1 and 30-40% of grade 2 tumors metastasize.

Our study has several limitations. It is a retrospective study and includes patients treated between 1974 and 2017, during which time diagnostic and treatment modalities have changed dramatically. The study population is a selected group of patients who received radiation, and some patients were referred for specialized radiation at a tertiary care hospital, which could affect patterns of disease and progression. The small sample size limited the number of confounders included in multivariable analysis. Pathology slides were not retrospectively re-reviewed for this study. Due to the inclusion of patients treated over multiple decades, pathologic data such as mitotic rate, Ki-67 index, and immunohistochemical markers for NETs were inconsistently reported. Additionally, the classification systems for NETs have changed over time and other factors such as hormone status (functional versus non-functional) were not consistently reported. While our cases were collected from a prospectively maintained database with histologically confirmed grade 1-2 NETs, these limitations introduce risk for misclassification bias. As described in our methods, where possible, the 2017 World Health Organization classification system was used in this paper.

Overall, this study reports a relatively large series of patients with grade 1-2 NETs treated with predominantly modern external beam RT. Consistent with previous small single institution retrospective studies, we show that grade 1-2 NETs can respond to RT. RT should be considered in the management of patients with grade 1-2 NETs, particularly in the setting of unresectable or metastatic disease.

## Data Availability

Data from this study are not being made publicly available.

